# Clinical implications of rare and common variation in preimplantation genetic testing for breast cancer

**DOI:** 10.1101/2025.10.08.25337603

**Authors:** Todd Lencz, Upasana Bhattacharyya, Liraz Klausner, Jibin John, Shai Carmi

## Abstract

Recently, some clinics have begun using preimplantation genetic testing for monogenic disorders (PGT-M) for moderately penetrant breast cancer (BC) risk variants, while other clinics use polygenic risk scores (PRS) in the context of preimplantation embryo screening. Using both simulation and formal mathematical approaches, we evaluated: 1) in what circumstances embryo selection using PRS could lead to systematically erroneous results due to failure to consider monogenic carrier status; and 2) whether PGT-M for moderate penetrance variants could lead to erroneous results due to unassessed, yet elevated PRS. Variants in *BRCA1, BRCA2*, and *PALB2* resulted in a risk distribution that was essentially disjoint from the non-carriers, regardless of PRS. By contrast, for moderately penetrant genes, standard PGT-M would fail to select the lowest risk embryo approximately 5% of the time due to elevated PRS. This complex interplay suggests that caution should be exercised when considering preimplantation genetic testing involving exclusively monogenic variants of moderate penetrance or polygenic scores.

## Introduction

Preimplantation genetic testing for monogenic disorders (PGT-M) has traditionally been used to avoid transmission of alleles with (virtually) complete penetrance for severe, early-onset disease. However, the use of PGT-M for variably penetrant variants for adult-onset diseases, such as breast cancer (BC), has become increasingly adopted by clinics^1^ and accepted by ethicists^2^. While the BC variants that are most commonly screened in PGT-M are highly penetrant rare variants in *BRCA1/2* (odds ratios ranging from 5-10^3,4^), PGT-M is now also performed for rare protein-truncating variants (PTVs) in genes such as *CHEK2, ATM, BARD1, RAD51C*, and *RAD51D*^5–7^. These have been labeled as genes of “moderate penetrance,” with odds ratios (ORs) for BC for PTV heterozygotes in the range of 1.5-2.5^3,4^.

Separately, polygenic risks scores (PRS) have been developed in recent years to predict polygenic disease risk by combining hundreds of alleles of small effect; a landmark study demonstrated that high (top decile) BC PRS conveys ORs similar to those of moderate penetrance mutations in monogenic risk genes^8^. Moreover, evidence from large-scale prospective cohorts of adult women has demonstrated that the penetrance of BC rare variants is modified by an individual’s PRS^9–12^. Importantly, the effect of PRS on BC risk in rare variant heterozygotes is additive on the liability scale, and effect sizes are generally consistent across the entire range of variants and PRS^13^.

The use of PRS to screen IVF embryos for future risk of complex disease has been available in the United States and other countries for more than five years^14^. It has been suggested that the utilization of such polygenic embryo screening (PES) for BC without concomitant examination of *BRCA1* may lead to inaccurate classification of embryos and subsequent implantation of a high-risk embryo^15^. Specifically, it was shown that the effect size of *BRCA1* on BC risk is so large that carriers had greater risk than all non-carriers, regardless of PRS^15^. However, to date, there has been no examination of other rare BC risk variants in the context of PES; conversely, the role of PRS in the context of PGT-M for BC has not been studied at all.

The present study was designed as a statistical analysis, using both simulation and formal mathematical approaches, to examine the relative contribution of BC rare variants and PRS in the setting of preimplantation genetic testing. Specifically, we sought to determine: 1) whether rare variants in genes other than *BRCA1* were sufficiently penetrant to lead to systematically inaccurate prioritization of embryos assessed using PES without PGT-M; and 2) whether polygenic risk was ever sufficient to yield incorrect prioritization of embryos assessed using PGT-M without PES.

## Results

### Generation of risk estimates

Consistent with previously reported results^15^, *BRCA1* carriers (**Figure 1**, red curve), are estimated to have greater lifetime BC risk (through age 80) than non-carriers (blue curve) at any PRS. Specifically, *BRCA1* carriers with the lowest PRS considered here (z-score=-3) have higher risk even compared to non-carriers with the highest PRS (z=+3; dashed horizontal red line). *BRCA2* (brown) and *PALB2* (pink) carriers demonstrate a small overlap with the non-carrier risk distribution at the extremes. As indicated by the dashed blue line in Figure 1, carriers of *BRCA2* would need to be in the bottom ∼1^st^ percentile for PRS (Z<-2.3) to have approximately equivalent risk to non-carriers with PRS in the top ∼1^st^ percentile of the population (Z>2.3).

**Figure 1.**
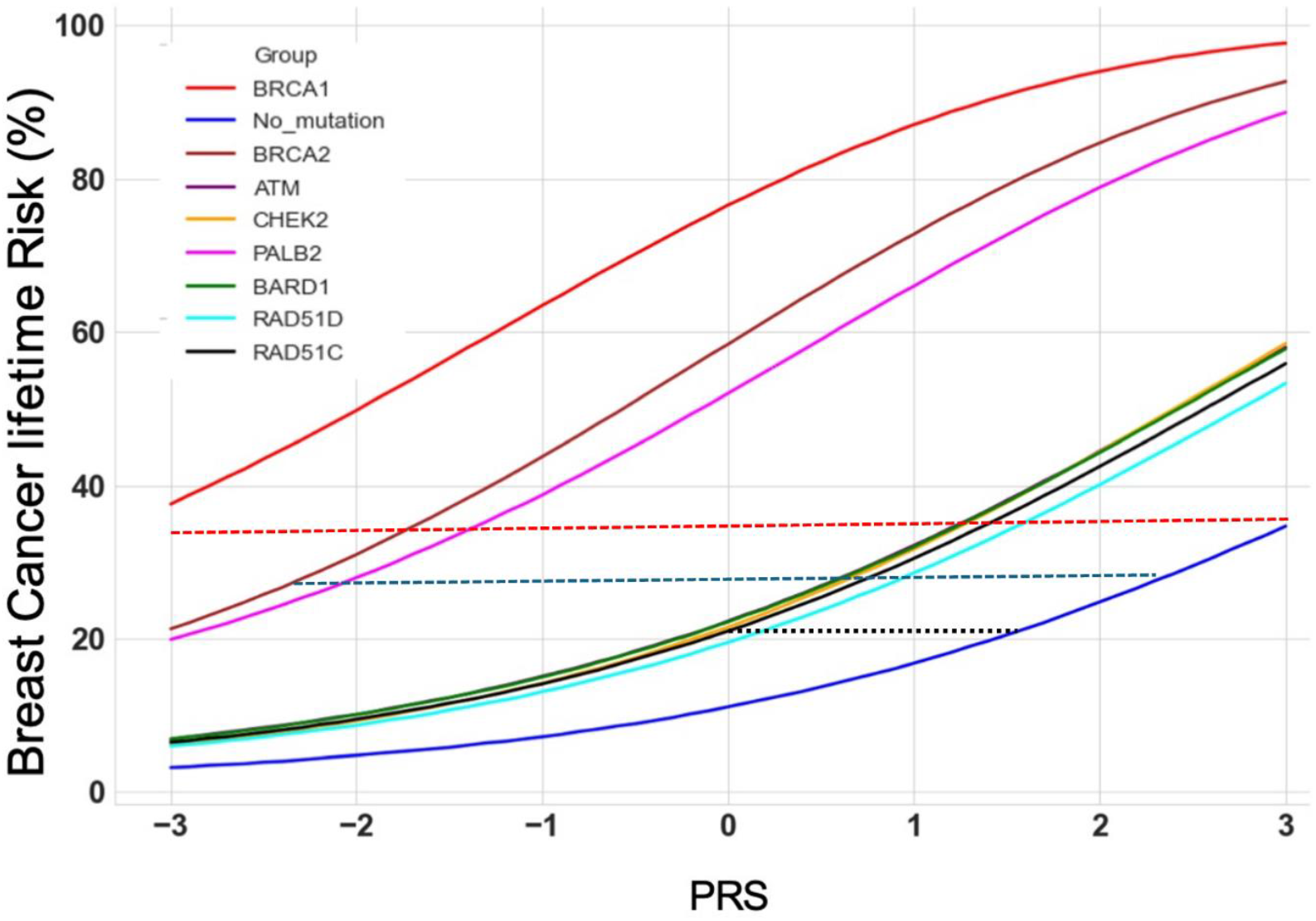
Lifetime risk for breast cancer (y-axis; percentages) as a function of the polygenic risk score (PRS) z-score (x-axis), for individuals heterozygous for a rare variant in each breast cancer risk gene, as well as non-carriers.

### Embryo simulation and risk evaluation

As sibling embryos are extremely unlikely to have such disparate PRS (2 per million in our model), the *BRCA2* and *PALB2* distributions are essentially disjoint from non-carrier distributions in the context of polygenic embryo screening.

Hence, as with *BRCA1*^15^, any embryo carrying an un-assessed pathogenic *BRCA2* or *PALB2* variant would be at elevated risk if selected on the basis of polygenic score alone.

By contrast, variants in several moderately penetrant genes confer lifetime risk levels that overlap the risk of many non-carriers (**Figure 1**). Specifically, the risk due to these variants (OR∼2) is approximately equivalent to a 1.5 standard deviation (SD) increase in PRS (black dotted line). Thus, in the context of PGT-M for moderate penetrance BC genes, a non-carrier embryo with a PRS more than ∼1.5 SD higher than a carrier embryo in the same cycle would have greater genetic liability to BC. Simulations demonstrated that, if there are two female euploid embryos available for implantation, a >1.5 SD difference in PRS would be observed 13.4% of the time. In half of these (6.7%), the embryo with the higher PRS would be the non-carrier embryo, and standard PGT-M would result in selection of the “wrong” (higher risk) embryo. **Figure 2** generalizes this for any number of (female) embryos, showing the proportion of cycles where the lowest risk embryo is a rare variant carrier, given at least one carrier and one non-carrier embryo in the batch. In realistic settings, in which the number of available embryos is small, standard PGT-M would fail to select the lowest risk embryo approximately 5% of the time. Essentially identical results can be derived analytically (see **Appendix**).

**Figure 2.**
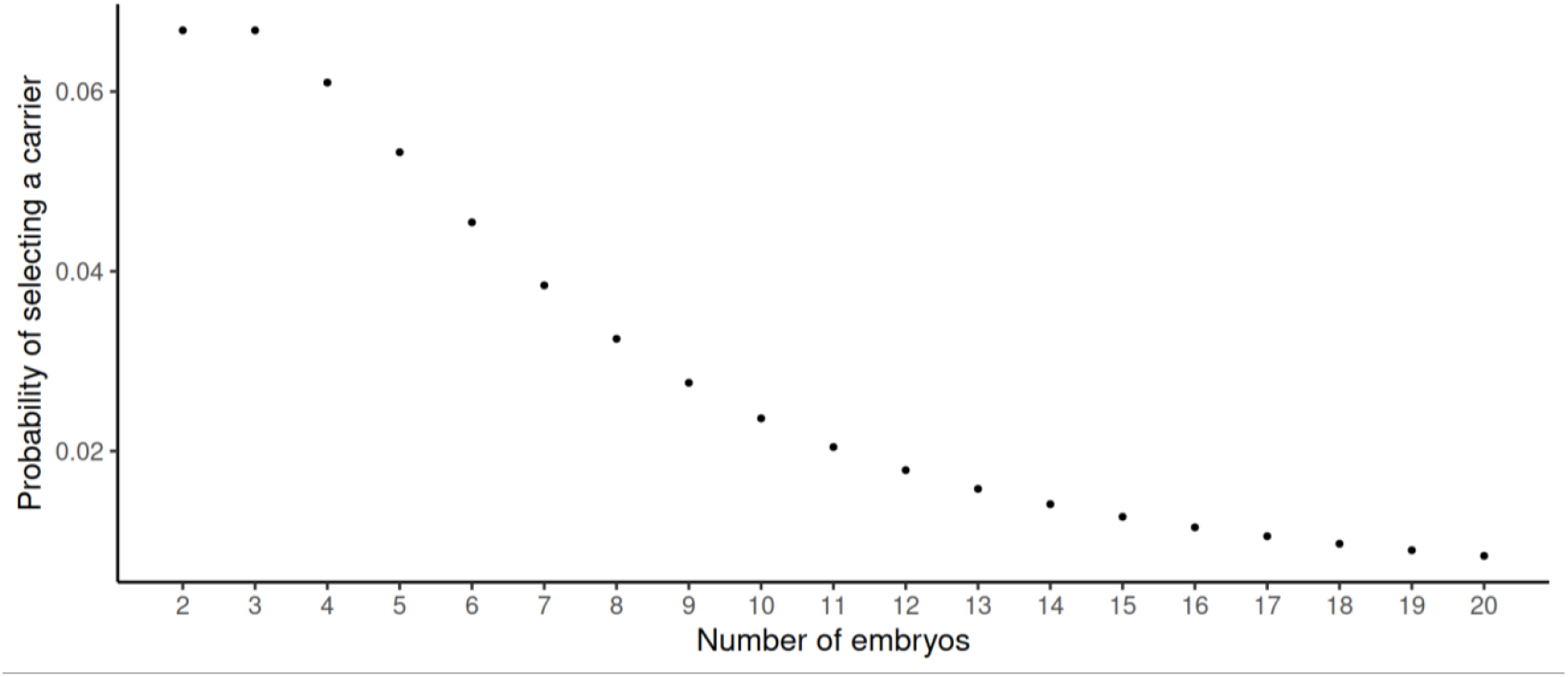
The proportion of cycles where the lowest risk embryo is a rare variant carrier, given at least one carrier and one non-carrier embryo in the batch.

## Discussion

The present study has two main findings, each with clinical implications for the conduct of preimplantation genetic testing. First, we demonstrated that BC risk for carriers of *BRCA2* and *PALB2* is higher than that for non-carriers across ∼98% of the range of possible values of PRS. In offspring of carriers, polygenic embryo screening that does not take such variants into account (as with *BRCA1*^15^) would have ∼50% chance of selecting an embryo at elevated risk. Currently, providers of PES have differing practices with respect to the assessment of family history, population risk factors (e.g., Ashkenazi Jewish heritage), and screening for penetrant rare variants^14^, the present results strongly suggest that careful consideration of these risk factors is important if cancer risk reduction is desired.

Second, we observed that embryo selection using PGT-M for carriers of “moderate penetrance” BC risk variants would select the “wrong” embryo ∼5% of the time (assuming a female child is desired and that both carrier and non-carrier embryos are available). PGT-M for these genes is not yet commonplace, but has been reported by multiple clinics^5–7^ and may become more common as genetic testing increases in frequency of use and breadth of coverage^16^. Given its novelty, there may be a naïve assumption that PGT-M for moderate penetrance BC risk genes is comparable to the more longstanding (though previously controversial) practice of PGT-M for *BRCA1*. Relatedly, there is a common misconception that there is little variability in PRS across embryos within a given batch, since they share the same parents^14^; to the contrary, the present study shows that differences of 1.5 standard deviations or more are not rare.

Our results can be considered by clinicians in several ways. First, the results reinforce a recent statement by the American College of Medical Genetics and Genomics (ACMG)^17^ that “PGT-M for moderate penetrance genes, such as *CHEK2*, is controversial, and, in general, not recommended, considering that cancer risks associated with *CHEK2* are not fully defined and because cancer risk is significantly modified by other factors” (p. 17). Regarding PES (also known as PGT-P), the ACMG has recently concluded that^18^ “At this time, there is insufficient evidence for the clinical utility of PRS testing for embryo selection. It should not be offered as a clinical service.” (p. 11). However, if clinicians and/or patients consider utilization of PGT-M for moderate penetrance genes, practitioners may wish to consider incorporation of PRS to obtain quantitative risk estimates per embryo.

By contrast, carriers of high-penetrance BC variants such as those in *BRCA1/2* would not benefit from PES if there is only a single non-carrier embryo; this embryo would necessarily have the lowest risk. However, *BRCA1/2* carriers could benefit incrementally from the integration of polygenic scores into the preimplantation genetic testing protocol, if there are two or more non-carrier embryos to choose from. Potential risk reductions amongst non-carrier embryos would follow the patterns we have previously described^25^, which can be examined on our publicly available risk calculator app (https://polygenicembryo.shinyapps.io/pestimate/)^19^. Such risk reductions would likely be small due to the limited number of non-carrier embryos from the batch. If only carrier embryos are available, and additional cycles are not possible or not desired, Figure 1 also demonstrates that polygenic risk scores impact lifetime BC risk in *BRCA1/2* carriers. However, such effects would probably not impact future medical recommendations for these offspring; regardless of polygenic risk score, all *BRCA1* carriers and ∼98% of *BRCA2* carriers have lifetime risk greater than any individuals with no rare risk variants and all would be regarded as high-risk for purposes of clinical management^20^.

More broadly, recent genetic evidence across multiple complex, adult-onset diseases suggests that the additive nature of rare and common variation is the rule, rather than the exception^13,21^. While breast cancer is the most well-studied disease in this regard^22,23^, recent studies have generated similar results in diseases ranging from Parkinson’s disease^24^ to chronic kidney disease^25^ to diabetes^26^. It is noteworthy that for each of these diseases, the effects of known monogenic rare variants were comparable to *BRCA1/2* in the present study; disease susceptibility is higher in carriers than in non-carriers at any level of PRS. Consequently, the implications for preimplantation genetic testing are limited to situations in which only carriers or only non-carriers are being compared with respect to PRS.

However, as sample sizes grow and association techniques improve, rare variants of moderate penetrance will be increasingly uncovered^27,28^, as are “core genes” with pleiotropic effects^29^. If preimplantation genetic testing were to expand beyond the most highly penetrant risk genes, it will become necessary to gain a greater understanding of the complex interplay between rare variants and polygenic risk scores. This interaction is most relevant for rare variants conferring odds ratios of ∼2; the moderate effect BC genes displayed in Figure 1 convey approximately 2-fold increase in risk, while *PALB2* has OR∼5^3,4^. At the same time, it should be acknowledged that unassessed rare variants do not lower the published accuracy of the polygenic risk scores at the population level; these already represent a portion of the unaccounted variance in any PRS model.

The complexity of these interactions further accentuates the need for appropriate genetic counseling in the context of preimplantation genetic testing for moderate penetrance and/or polygenic risk. Professionals working in assisted reproductive technologies have consistently voiced concerns that polygenic embryo screening may place undue strain on their capacity to provide adequate genetic counseling ^30–32^. Polygenic screening adds to the existing challenges in providing fully informed consent for preimplantation testing, given differing understandings of risk for patients relative to providers^31,33^, the potential implication of family members for cascade testing^34^, and the already overburdened genetic counseling workforce^35^.

The present study has several notable limitations. Most importantly, it relied upon statistical approaches, including simulation and mathematical modeling, rather than empirical data. However, such approaches are necessary, given that an 80-year clinical follow-up study is a practical impossibility^14^; it is noteworthy that PGT-M for *BRCA1* carriers has become relatively common in the absence of such evidence. The present study only considered BC risk, although several of the moderate penetrance genes also elevate risk for ovarian and other cancers.

Given that polygenic risk for breast and ovarian cancer are only moderately correlated^36^, the results presented in Figure 2 would be somewhat altered in the context of predicting total cancer risk. It is important to note that all of our calculations, and more generally, the possibility of preimplantation genetic testing for disease, is premised on the availability of sufficient embryos for testing; many patients with fertility issues will prioritize achieving a single live birth^14^.A final limitation is that, in Figure 2, we considered only selection amongst female embryos within batches with both carrier and non-carrier embryos; compared to females, males have extremely low risk for breast cancer at each level of rare and common variant risk^37,38^. A more complex framework encompassing alternative scenarios will depend on parental reproductive goals and choices (e.g., regarding embryo sex and maximal number of IVF cycles) and will remain for future development.

## Methods

### Generation of risk estimates

To compare risks across carrier and non-carrier embryos, we first generated risk estimates for each known BC risk gene across the range of PRS z-scores (-3 ≤ Z ≤ 3). For this step, we utilized the CanRisk Tool^39^, which calculates the additive risk based on the BOADICEA model^9^, incorporating data from the largest available BC genetic datasets. To generate lifetime risk estimates from the CanRisk Tool, we set date of birth to the earliest available value (resulting in an estimate for age 80y) and sex to female, with all other demographic and medical variables left blank or set to “unknown”. We then performed API calls to estimate BC risk across the range of polygenic risk scores (in z-score increments of 0.1), calculated using the well-validated set of 313 SNPs from the Breast Cancer Association Consortium (BCAC)^40^. We then repeated this procedure, with all settings held constant, except for presence of a single known risk variant in one of the genes available in the CanRisk Tool: *BRCA1, BRCA2, PALB2, CHEK2, ATM, BARD1, RAD51C*, or *RAD51D*. Results are plotted in **Figure 1**, which displays the lifetime risk for breast cancer (y-axis; percentages) as a function of the PRS z-score (x-axis), for individuals heterozygous for a rare variant in each gene, as well as non-carriers.

### Embryo simulation and risk evaluation

To simulate the risk of embryos, we used the liability threshold model (LTM)^41^, which is commonly utilized in genetic modeling (including PES^42–44^) and empirical studies of real-world populations^45^. The LTM posits that the risk for a given complex disease is a function of a normally distributed latent continuous trait, the “liability”, which represents a sum of genetic and non-genetic risk factors. An individual is “affected” if the overall liability exceeds a certain threshold. In our study, the liability is a sum of an embryo’s PRS, risk due to a pathogenic variant, and non-genetic risk factors.

We simulated the PRS for each embryo in a batch of *n* embryos as a sum of two factors: the shared component across all embryos in the batch, equal to the average parental PRS; and an embryo-specific component (due to Mendelian segregation of all alleles included in the PRS)^42,43^. Parental PRS values are drawn from a normal distribution with variance equal to *r*^2^, the proportion of variance in liability across the population explained by the PRS. The embryo-specific component is also normally distributed, with half the population variance^46^. To simulate the risk due to a pathogenic variant, we assumed that one of the parents is heterozygous for a dominant pathogenic variant, such that each embryo inherits the variant with probability=0.5.

The overall liability combines the genetic risk with a random, normally distributed, value of non-genetic (environmental and other) liability. Given that at conception only genetic risks are known, the probability of disease (or risk) of each embryo is set to the probability that the future non-genetic risk will push the liability above the disease threshold.^41^

We performed 100,000 simulations for each value of *n* female embryos, where *n* ranged from 2-20. To specifically study the interplay between PES and PGT-M, we assumed that at least one carrier and one non-carrier female embryo are available, and that a female embryo is desired. Further details, including an analytic solution that produced identical results to the simulation approach, are provided in the **Appendix**.

We used the results of these simulations to evaluate: 1) whether an embryo selected on the basis of having the lowest polygenic risk^42^ could be systematically (i.e., at all values of PRS) overridden by carrier status^15^ at a given risk gene; and 2) conversely, whether a non-carrier embryo, selected on the basis of PGT-M, might have higherrisk compared to a carrier embryo once polygenic risk is also evaluated.

## Data Availability

Data and code used in this study will be made available upon reasonable request.

## Data/Code Availability

Data and code used in this study will be made available upon reasonable request.

## Acknowledgments

Research reported in this publication was supported by the National Human Genome Research Institute of the National Institutes of Health under award number R01HG011711. The content is solely the responsibility of the authors and does not necessarily represent the official views of the National Institutes of Health.

## Author Contributions

T.L. initiated the project, T.L. and S.C. conceptualized the article, T.L. and U.B, reviewed the literature, U.B., J.J., S.C., and L.K. performed the statistical simulations, T.L. and S.C. obtained the funding. The writing was performed by T.L. All authors were involved in discussions and participated in reviewing and editing the article and all approved the final version.

## Competing Interests

S.C. is a paid consultant and owns stock options at MyHeritage; he further declares an (unpaid) collaboration on specific projects with Juno Genetics. The other authors declare no conflicts of interest.

