## Appendix for "Clinical implications of rare and common variation in preimplantation genetic testing for breast cancer"

### The probability of a pathogenic variant carrier embryo to have the lowest overall risk

#### 1 Model

The model is based on [1]. There are $n$ embryos. For $i = 1, \ldots, n$,

$$\begin{aligned} \text{Carrier}_i &\sim \text{Ber}(0.5) \\ \text{PRS}_i &= x_i + c \\ c &\sim N(0, 0.5) \\ x_i &\sim N(0, 0.5) \\ s_i &= \text{PRS}_i + \alpha \text{Carrier}_i. \end{aligned}$$

We assume a dominant pathogenic variant and Mendelian transmission, such that each embryo is a carrier with probability 0.5. Next, $c$ is the shared component of the PRS, equal to the average parental PRS, and $x_i$ is the embryo specific component (due to Mendelian segregation). Carrying the pathogenic variant is assumed to increase the PRS by $\alpha$, such that $s_i$ is the final "combined" risk score.

#### 2 A fixed number of carrier and non-carrier embryos

In this section, we assume that the number of carrier and non-carrier embryos is fixed and known, and we derive the probability that the embryo with the minimal combined risk score is a carrier. Equivalently, it is the probability that the minimum risk score among non-carriers exceeds that of carriers. Assume that we have $k$ carrier and $n - k$ non-carrier embryos. Define $t$ as the minimum combined risk score among carriers and $s$ as the minimum score among non-carriers,

$$t \equiv \min_{i \in \text{carriers}} s_i = \min_{i \in \text{carriers}} \text{PRS}_i + \alpha = \min_{i \in \text{carriers}} x_i + c + \alpha \tag{1}$$

$$s \equiv \min_{i \in \text{non carriers}} \text{PRS}_i = \min_{i \in \text{non carriers}} x_i + c \tag{2}$$

The probability that the embryo with the minimum risk is a carrier (given $k$ carrier and $n-k$ non-carrier embryos) is then $P(s > t \mid k, n-k)$.

Denote $s_x = \min_{i \in \text{non carriers}} x_i$ and $t_x = \min_{i \in \text{carriers}} x_i$. The probability that the embryo with the minimal risk is a carrier is

$$P(s > t \mid k, n-k) = P\left(s_x > t_x + \alpha\right). \tag{3}$$

Using the law of total probability,

$$P(s_x > t_x + \alpha) = \int_{-\infty}^{\infty} P(s_x > t_x + \alpha \mid t_x) f(t_x) dt_x. \tag{4}$$

In Eq. (4), $f(t_x)$ is the density of $t_x$. It is the density of the minimum of $k$ independent random variables, each distributed normally with zero mean and variance $1/2$. The term $P(s_x > t_x + \alpha \mid t_x)$ is the one minus the cumulative distribution function of the minimum of $n-k$ independent random variables, again each normally distributed with zero mean and variance $1/2$. Thus,

$$f(t_x) = \sqrt{2}k\ \phi\left(\frac{t_x}{1/\sqrt{2}}\right)\left[1 - \Phi\left(\frac{t_x}{1/\sqrt{2}}\right)\right]^{k-1}, \tag{5}$$

and

$$P(s_x > t_x + \alpha \mid t_x) = \left[1 - \Phi\left(\frac{t_x + \alpha}{1/\sqrt{2}}\right)\right]^{n-k} \tag{6}$$

where $\phi(\cdot)$ and $\Phi(\cdot)$ are the probability density function (PDF) and cumulative distribution function (CDF), respectively, of the standard normal variable.

Therefore, the probability that the embryo with the minimal risk is a carrier is

$$\begin{aligned}
&P(s > t \mid k, n-k) = \\
&= \int_{-\infty}^{\infty} \left[1 - \Phi\left(\frac{t_x + \alpha}{1/\sqrt{2}}\right)\right]^{n-k} \sqrt{2}k\ \phi\left(\frac{t_x}{1/\sqrt{2}}\right)\left[1 - \Phi\left(\frac{t_x}{1/\sqrt{2}}\right)\right]^{k-1} dt_x \\
&= \sqrt{2}k \int_{-\infty}^{\infty} \phi\left(\frac{t_x}{1/\sqrt{2}}\right)\left[1 - \Phi\left(\frac{t_x}{1/\sqrt{2}}\right)\right]^{k-1}\left[1 - \Phi\left(\frac{\alpha + t_x}{1/\sqrt{2}}\right)\right]^{n-k} dt_x \\
&= k \int_{-\infty}^{\infty} \phi(z)\left[1 - \Phi(z)\right]^{k-1}\left[1 - \Phi\left(\sqrt{2}\left(\alpha + z/\sqrt{2}\right)\right)\right]^{n-k} dz \\
&= k \int_{-\infty}^{\infty} \phi(z)\left[1 - \Phi(z)\right]^{k-1}\left[1 - \Phi\left(z + \sqrt{2}\alpha\right)\right]^{n-k} dz.
\end{aligned} \tag{7}$$

#### 3 A fixed number of embryos

Here, we assume that the number of embryos $n$ is fixed, but the number of carrier and non-carrier embryos is random. Denote the number of carrier embryos as $n_c$, which has a binomial distribution, $n_c \sim \text{Bin}(n, 0.5)$. The probability of the embryo with the minimal risk score to be a carrier is

$$\begin{aligned}
P(s > t) &= \sum_{k=0}^{n} P(s > t \mid k, n-k) P(n_c = k) \\
&= \sum_{k=0}^{n} P(s > t \mid k, n-k) \binom{n}{k} \left(\frac{1}{2}\right)^n \\
&= \left(\frac{1}{2}\right)^n \sum_{k=0}^{n} \binom{n}{k} k \int_{-\infty}^{\infty} \phi(z) \left[1 - \Phi(z)\right]^{k-1} \left[1 - \Phi\left(z + \sqrt{2}\alpha\right)\right]^{n-k} dz \\
&= \left(\frac{1}{2}\right)^n \int_{-\infty}^{\infty} \phi(z) \sum_{k=0}^{n} \binom{n}{k} k \left[1 - \Phi(z)\right]^{k-1} \left[1 - \Phi\left(z + \sqrt{2}\alpha\right)\right]^{n-k} dz \\
&= n \left(\frac{1}{2}\right)^n \int_{-\infty}^{\infty} \phi(z) \left[2 - \Phi(z) - \Phi\left(z + \sqrt{2}\alpha\right)\right]^{n-1} dz \qquad (8)
\end{aligned}$$

For the last step, we used the following identity,

$$\sum_{k=0}^{n} \binom{n}{k} k x^{k-1} y^{n-k} = n(x+y)^{n-1}. \qquad (9)$$

To prove the identity, we start with the binomial theorem

$$(x+y)^n = \sum_{k=0}^{n} \binom{n}{k} x^k y^{n-k} \qquad (10)$$

and take the derivative of both sides with respect to $x$.

#### 4 Having at least one carrier and one non-carrier embryos

If all embryos are carriers or all embryos are non-carriers, the carrier status of the minimal risk embryo is of less interest. Therefore, in this section, we consider the case when we have at least one carrier embryo and at least one non-carrier embryo, i.e., $1 \leqslant n_c \leqslant n-1$. The probability distribution of the number of carrier embryos is simply the truncated binomal,

$$P(n_c = k \mid 1 \leqslant n_c \leqslant n-1) = \frac{P(n_c = k)}{1 - P(n_c = 0) - P(n_c = n)} = \frac{\binom{n}{k}\left(\frac{1}{2}\right)^n}{1 - 2 \cdot (1/2)^n}. \qquad (11)$$

In this setting, the probability that the embryo with the minimal risk is a carrier is

$$
\begin{aligned}
&P(s > t \mid 1 \leqslant n_c \leqslant n-1) = \\
&= \sum_{k=1}^{n-1} P(s > t \mid k, n-k) P(n_c = k \mid 1 \leqslant n_c \leqslant n-1) \\
&= \sum_{k=1}^{n-1} P(s > t \mid k, n-k) \frac{\binom{n}{k}\left(\frac{1}{2}\right)^n}{1-2^{1-n}} \\
&= \frac{1}{2\left(2^{n-1}-1\right)} \sum_{k=1}^{n-1} \binom{n}{k} P(s > t \mid k, n-k) \\
&= \frac{1}{2\left(2^{n-1}-1\right)} \sum_{k=1}^{n-1} \binom{n}{k} k \int_{-\infty}^{\infty} \phi(z) \left[1-\Phi(z)\right]^{k-1} \left[1-\Phi\left(z+\sqrt{2}\alpha\right)\right]^{n-k} dz \\
&= \frac{1}{2\left(2^{n-1}-1\right)} \int_{-\infty}^{\infty} \phi(z) \sum_{k=1}^{n-1} \binom{n}{k} k \left[1-\Phi(z)\right]^{k-1} \left[1-\Phi\left(z+\sqrt{2}\alpha\right)\right]^{n-k} dz \\
&= \frac{n}{2\left(2^{n-1}-1\right)} \int_{-\infty}^{\infty} \phi(z) \left\{\left[2-\Phi(z)-\Phi\left(z+\sqrt{2}\alpha\right)\right]^{n-1} - \left[1-\Phi(z)\right]^{n-1}\right\} dz.
\end{aligned} \tag{12}
$$

For the last step, we used the following identity,

$$\sum_{k=1}^{n-1} \binom{n}{k} k x^{k-1} y^{n-k} = n\left[(x+y)^{n-1} - x^{n-1}\right]. \tag{13}$$

This identity can be derived from that of Eq. (9), by noting that the term corresponding to $k = 0$ is zero and then extending the sum to $k = n$ and subtracting the last term,

$$
\begin{aligned}
\sum_{k=1}^{n-1} \binom{n}{k} k x^{k-1} y^{n-k} &= \sum_{k=0}^{n} \binom{n}{k} k x^{k-1} y^{n-k} - n x^{n-1} \\
&= n\left[(x+y)^{n-1} - x^{n-1}\right].
\end{aligned} \tag{14}
$$

### 5 Numerical evaluation

To generate the figure in the main paper, we evaluated the integrals in Eq. (12) in `R` using the function `integrate`. We validated the solution to Eq. (12) using simulations from the model of Section 1.
